# The amino acid stress sensing eIF2α kinase GCN2 is a survival biomarker for malignant mesothelioma

**DOI:** 10.1101/2023.03.24.23287516

**Authors:** Lyssa T. Gold, Susan E. Bray, Neil M. Kernohan, Nina Henderson, Maisie Nowicki, Glenn R. Masson

## Abstract

**Background:** Malignant mesothelioma is a tumour that is strongly associated with a history of asbestos exposure and which derives from mesothelial cells that line the serous cavities of the body. The tumour most commonly arises in the pleural cavity, but can also arise in the pericardium, peritoneum and tunica vaginalis. At present the lesion has a very poor prognosis and is an incurable form of cancer with median survival times of up to 19 months being quoted for some histological subtypes. A large proportion of mesotheliomas have been shown to be arginine auxotrophic, leading to new research for therapeutics which might exploit this potential vulnerability.

**Method:** We measured the levels of General Control Non-derepressible 2 (GCN2) protein in malignant mesothelioma tumour samples and determined whether these levels correlate with clinical outcomes.

**Results:** We observed that the expression levels of GCN2 correlated with patient survival and was an independent prognostic variable in pairwise comparison comparisons with all available clinical data.

**Conclusion:** These findings suggest that GCN2 levels provides prognostic information and may allow for stratification of care pathways. It may suggest that targeting GCN2 is a viable strategy for mesothelioma therapy development.

## Background

Malignant mesothelioma (MM) is a treatment-limited cancer derived from mesothelial cells that form the surface level of the protective serous membrane coating the serous cavities of the body, namely, the pleural, pericardial and peritoneal cavities as well as the tunica vaginalis, an embryological invagination of the peritoneum into the scrotum that encloses the testis and paratesticular tissues. (1). Asbestos exposure is the predominant cause of MM(1), whilst the majority of cases (90%) occur in the pleura, with the - peritoneum lining the abdominopelvic cavity contributing 7%-10% of MM cases (2). Without treatment, median survival is 7-10 months, reaching 19 months for some histological subtypes(3), which increases to a maximum of 22 months with treatment (4). The most effective chemotherapeutic strategy for MM is a combinatorial treatment using cisplatin-pemetrexed, however this yields a poor response rate of only 41.3% (5).This, coupled with a five-year survival rate of only 5% (6), underscores the need for greater understanding of the cellular processes underpinning MM pathogenesis to develop more effective, targeted, therapies and improve patient outlook.

Recent randomised clinical trials have demonstrated an increased survival benefit for immune checkpoint inhibitors (7) and arginine depletion strategies(8). One potential avenue for therapeutic development may be found in the observation that around 60% of malignant pleural mesothelioma tumours (which represents ∼90% of all MM) do not express asparagine synthetase (ASS1) (9). ASS1 catalyses the rate limiting step in arginine biosynthesis, and its loss, a common occurrence in multiple cancers, makes tumours reliant on extracellular arginine to survive (10). The arginine auxotrophic phenotype is currently being investigated therapeutically through administration of a pegylated arginine deiminase (ADI-PEG-20) in the Arginine Deiminase and Mesothelioma (ADAM) study (8), along with the standard combination of pemetrexed (Pem) and cisplatin (11).

Analogously, the molecular basis of ADI-PEG-20 treatment has been demonstrated to depend on the GCN2-eIF2α-ATF4 pathway in arginine-dependent invasive bladder cancer (12). General Control Nonderepressible 2 (GCN2) is an protein kinase which is activated through amino-acid starvation stress which has been implicated in multiple cancers (13,14). GCN2 activation may be dependent on ribosomal stalling (15,16), deacylated tRNA levels increasing (17,18), or P-stalk proteins associating with the ribosome (17,19,20), all of which result in autophosphorylation and subsequent phosphorylation of the S51 site of the eukaryotic initiation factor eIF2α which prevents the formation of translational pre-initiation complexes. This results in the Integrated Stress Response (ISR) – a global repression of mRNA translation and the upregulation of the transcription factor Activating Transcription Factor 4 (ATF4) – a transcription factor which promotes cell survival, unless under chronic or acute stress levels, where instead apoptosis is activated (13). There are ongoing trials GCN2-targetting therapeutics (21) and several pre-clinical inhibitors in development (22–24).

ASS-1 deficient bladder cancer cells are ADI-PEG-20 sensitive, activating the GCN2-eIF2α-ATF4 pathway and inducing C/EBP homologous protein (CHOP) expression. This resulted in the selective apoptosis in the ASS1-deficient bladder cancer cells. Furthermore, it has been previously noted through the work of Dalton *et al*. that CHOP expression is a biomarker of malignant mesothelioma (25). We hypothesised that GCN2 levels would be a higher in biopsy samples taken from cases of malignant mesothelioma and may be a prognostic biomarker for patient outcomes. We measured GCN2 levels in formalin-fixed paraffin-embedded samples using immune-histochemical means. Here we report that GCN2 levels are a strong prognostic marker for mesothelioma patient outcomes.

## Methods

### Tissue microarrays

Tissue microarrays (TMA’s) were obtained from Mesobank tissue bank based at the NHS Royal Papworth Hospital, the largest UK pleural mesothelioma biobank(26). TMA’s were prepared by the biobank for 204 patients with cores of 0.6 mm diameter, with between four and five replicates for each patient included on concurrent locations within the set. Demographic data was provided for each patient, including age at diagnosis, time to death, gender, TNM classification, and histopathology (table 1). Normal kidney, heart, and liver tissue was dispersed throughout the slide sets for the purpose of microscope orientation, and these also underwent staining. Consistency in staining was observed for these tissues within and between slides, indicating that staining conditions were consistent and not a source of error.

**Table 1:** TMA Cohort demographics.

|  | Number of patients | Total number of patients | % |
| --- | --- | --- | --- |
| **Gender** |  | 204 |  |
| Male | 184 |  | 90.2 |
| Female | 20 |  | 9.8 |
| **Histology** |  | 204 |  |
| Epithelioid | 126 |  | 61.8 |
| Biphasic | 47 |  | 23.0 |
| Sarcomatoid | 29 |  | 14.2 |
| Desmoplastic | 2 |  | 1.0 |
| **Alive** |  | 201 |  |
| Yes | 4 |  | 1.9 |
| No | 197 |  | 98.0 |
| **Tumour extent (T)** |  | 65 |  |
| 0 | 1 |  | 1.5 |
| 1/1b | 8 |  | 12.3 |
| 2 | 19 |  | 29.2 |
| 3 | 20 |  | 30.8 |
| 4 | 17 |  | 26.2 |
| **Node spread (N)** |  | 63 |  |
| 0 | 37 |  | 58.7 |
| 1 | 8 |  | 12.7 |
| 2 | 16 |  | 25.4 |
| 3 | 2 |  | 3.0 |
| **Metastasis** |  | 63 |  |
| 0 | 40 |  | 63.5 |
| 1/1a | 22 |  | 36.5 |

**Table 2:** Pairwise combinations including GCN2 Score.

| Paired comparison | HR(Exp(B)) | 95% CI low | 95% CI high | p-value | *n* |
| --- | --- | --- | --- | --- | --- |
| GCN2 Score | 0.5846 | 0.4246 | 0.8169 | 0.0013 | 194 |
| Histology | 1.961 | 1.442 | 2.653 | <0.001 |  |
| GCN2 Score | 1.285 | 0.6310 | 2.447 | 0.4643 | 59 |
| Disease State M | 2.333 | 1.272 | 4.215 | 0.0053 |  |

### Immunohistochemistry

Antigen retrieval and de-paraffinisation was performed using DAKO EnVision^™^ FLEX Target Retrieval solution (high pH) buffer in a DAKO PT Link. Sections were blocked in EnVision^™^ Flex Peroxidase-Blocking Reagent and incubated overnight at 4□ with anti-GCN2 rabbit monoclonal antibody (Invitrogen MA5-32704) at a dilution of 1-100.Immunostaining using DAKO EnVision^™^ FLEX system (Agilent Technologies) on a DAKO Autostainer Link48 was carried out according to manufacturer’s protocol. DAKO substrate working solution was used as a chromogenic agent for 2 × 5 mins and sections were counterstained with EnVision^™^ FLEX haematoxylin. Sections known to stain positively were included in each batch and negative controls were prepared by replacing the primary antibody with DAKO antibody diluent.

### Scoring

The methodology was based on previous mesothelioma immunohistochemical investigations (25). To quantify GCN2 levels, an image library compiling each tumour section was captured using LEICA ICC50 HD microscope at 10× magnification, using the Leica LAS EZ Application Suite Version 3.4.0. Individual sections were then scored on two scales. First, staining intensity scores from 0 to 5 were assigned independently by three persons and averaged. Second, proportion staining scores from 0 to 100, representing the percentage of all cells in the section stained. These scores were combined for each section to give a value from 0 to 500 and an average of all replicates calculated for each patient.

### Statistical analysis

Statistical analysis was carried out using GraphPad Prism 9 (GraphPad Prism version 9.5.1 for Windows, San Diego, California USA) and RStudio. Survival was assessed from date of histological diagnosis to death. Patients that were still alive were excluded from the survival analysis. Univariate survival analysis for demographic data and GCN2 scores used the log-rank (LR) test and was visualised as Kaplan-Meier (KM) survival curves, both in GraphPad Prism. The pROC package (27) in RStudio (28) was used to generate ROC (receiver operator characteristic) curves to determine whether GCN2 was a prognostic marker in malignant pleural mesothelioma (MPM) patients. C-statistics (also referred to as area under curve/AUC) were estimated by the package for logistic regression models fitted to one-year survival outcomes. Based on previous work in MPM (25,29), a significance cut-off of 0.65 for the C-statistic was used, where a value of 0.5 indicates a relationship governed by chance. Differences in GCN2 scores between histological subtypes (epithelioid, biphasic, sarcomatoid) were determined in GraphPad Prism using an ordinary one-way ANOVA with Bonferroni *post hoc* test to correct for family-wise error.

## Results

### Patient Cohort Characteristics

Five tissue microarray (TMA) slides were obtained from the MesobanK(26) containing malignant pleural mesothelioma (MPM) samples from a total of 204 patients. Accompanying patient demographic data are summarised in Table 1 and Figure 1. Clinical data were complete to January 2022. The majority of patients were male (90.2%) with a median age of 69 years. Epithelioid histology was the most prevalent (126 of 204, 61.8%), followed by biphasic (47 of 204, 23.0%), and sarcomatoid histological subtype (sarcomatoid | 29 of 204, 14.2%; desmoplastic | 2 of 204, 1.0%). TNM (tumour, node, metastasis) data was provided partially or entirely for 65 of 204 patients. For patients with this data available, 56.9% (T3-T4, 37 of 65) had more extensive tumours that covered all pleural surfaces in one side of the body. 41.3% showed some degree of lymph node involvement (N1-N3, 26 of 63), and 35.5% of cancers showed evidence of distant metastasis (M1/1a, 22 of 62). Four patients (1.9 %) were alive at time of data analysis, and the living status data were missing for three patients (1.5%). Three patients (1.5%) were excluded due to no tissue sections were present for these patients within the slide set.

**Figure 1:**
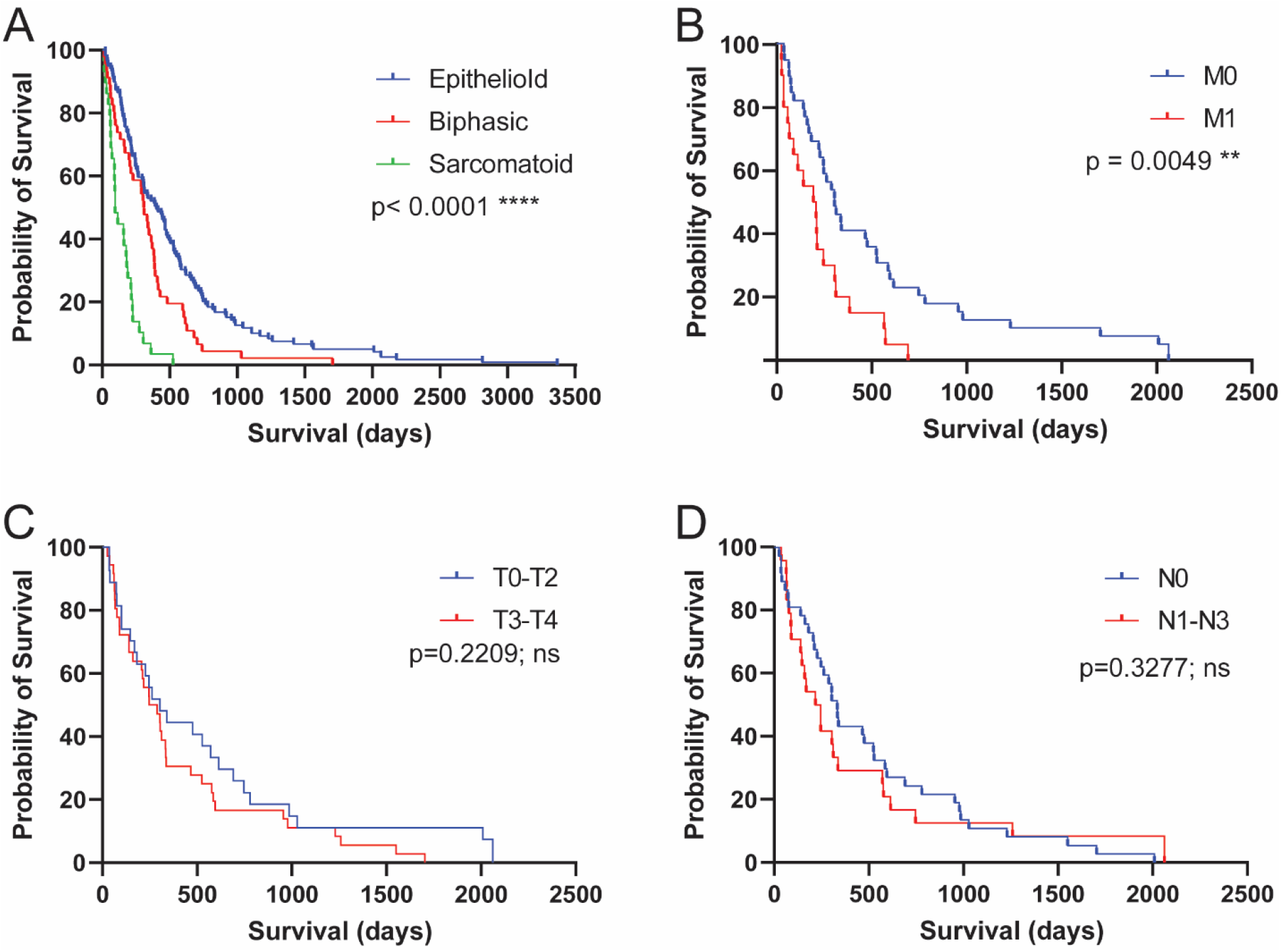
Mesothelioma TMA cohort properties and prognosis. Kaplan-Meier curves corresponding to patients captured in TMA demographic data. Statistical significance of figures A-D show in the top right, referring to Univariate survival analysis using the log-rank test. (**A**) Histological sub-type with epithelioid, biphasic and sarcomatoid. (**B**) Metastasis state?(**C**) Tumour score (**D**) Node score.

### Determination of GCN2 levels

The scoring of the TMA for GCN2 was performed in parallel by three persons (LTG, NH, and MN) all of whom where blinded to the clinical data. Intensity of GCN2 staining was scaled from 0 to 5 (0 for no discernible staining, 1 being little staining, 3 for moderate staining, 5 for intense staining) (See Figure 2A). Each section was then given a percentage value for the extent of malignant cells that were stained. The staining score for GCN2 levels was then calculated through multiplying the staining score with the extent percentage giving a range of 0 to 500 for each section. GCN2 staining showed no correlation with histological subtype, metastasis score or tumour score (See Figures 2B-D).

**Figure 2:**
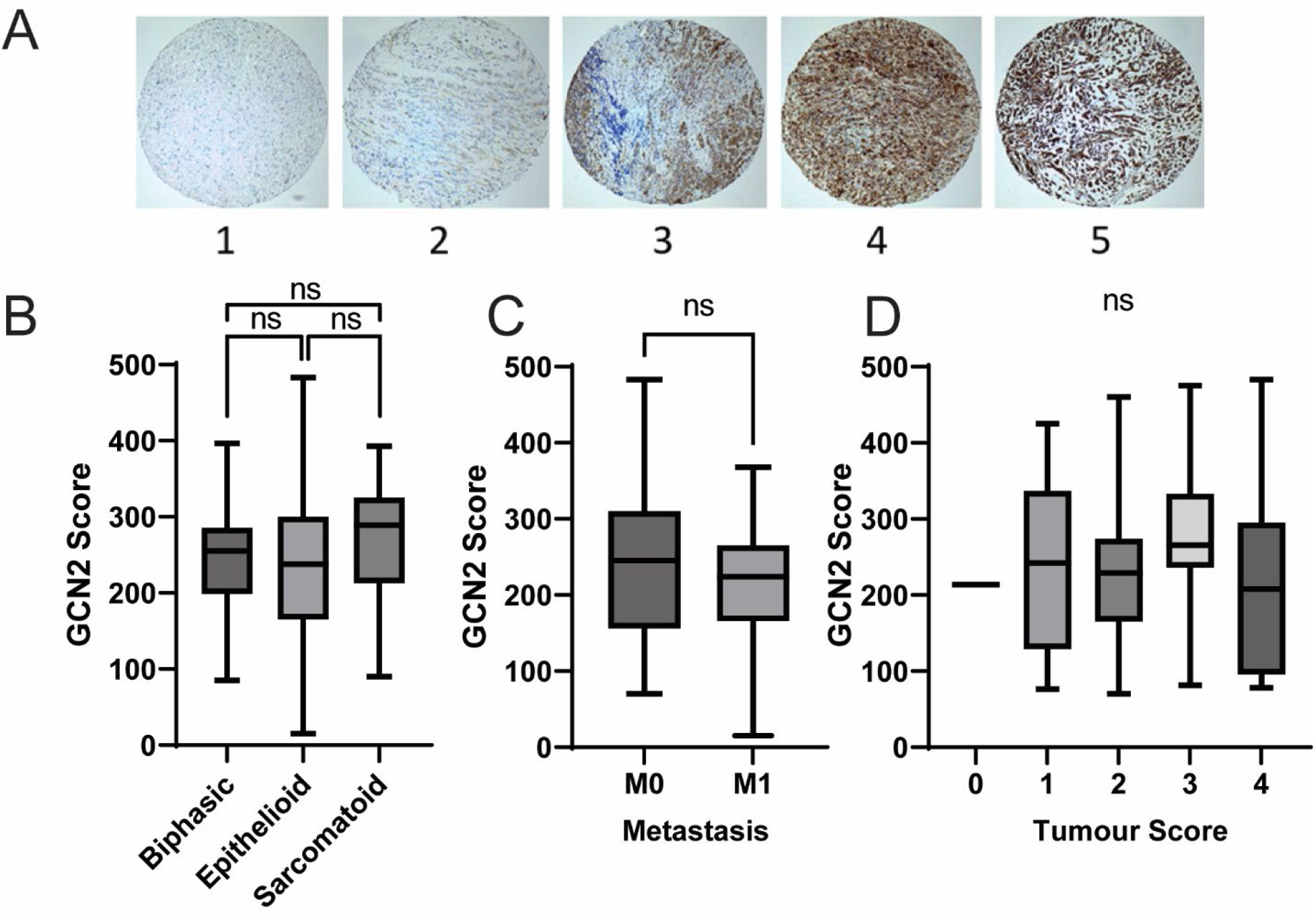
TMA staining for GCN2 characteristics. (**A**) Representative microarrays of intensity scores 1-5. (**B**) GCN2 Scores in different histological subtype. (**C**) GCN2 scores compared with metastasis scores. (**D**) GCN2 score in the different tumour score subtypes. Note there was only one instance of Tumour Score “0” in the dataset. In **B** and **D**, statistical differences were calculated by an ANOVA with Bonferroni *post-hoc* test, while for **C**, a non-paired t-test was used.

### Univariate Analysis

Histological subtype (p<0.0001) and metastasis scores (p = 0.0049) were prognostic with univariate analysis (see Figure 1), although neither tumour nor node score had any significant impact on survival. GCN2 scores (See Figure 3) had a significant association with prognosis (p=0.0009). Those cases scoring in the upper two quintiles (“High GCN2”) had a significantly shorter survival than those in the lower two quintiles (“Low GCN2”) or the rest of the cohort (“Remainder”) (High GCN2 -181.5 Days, Low GCN2 - 461.5 Days, Remainder 382.0 Days).

**Figure 3:**
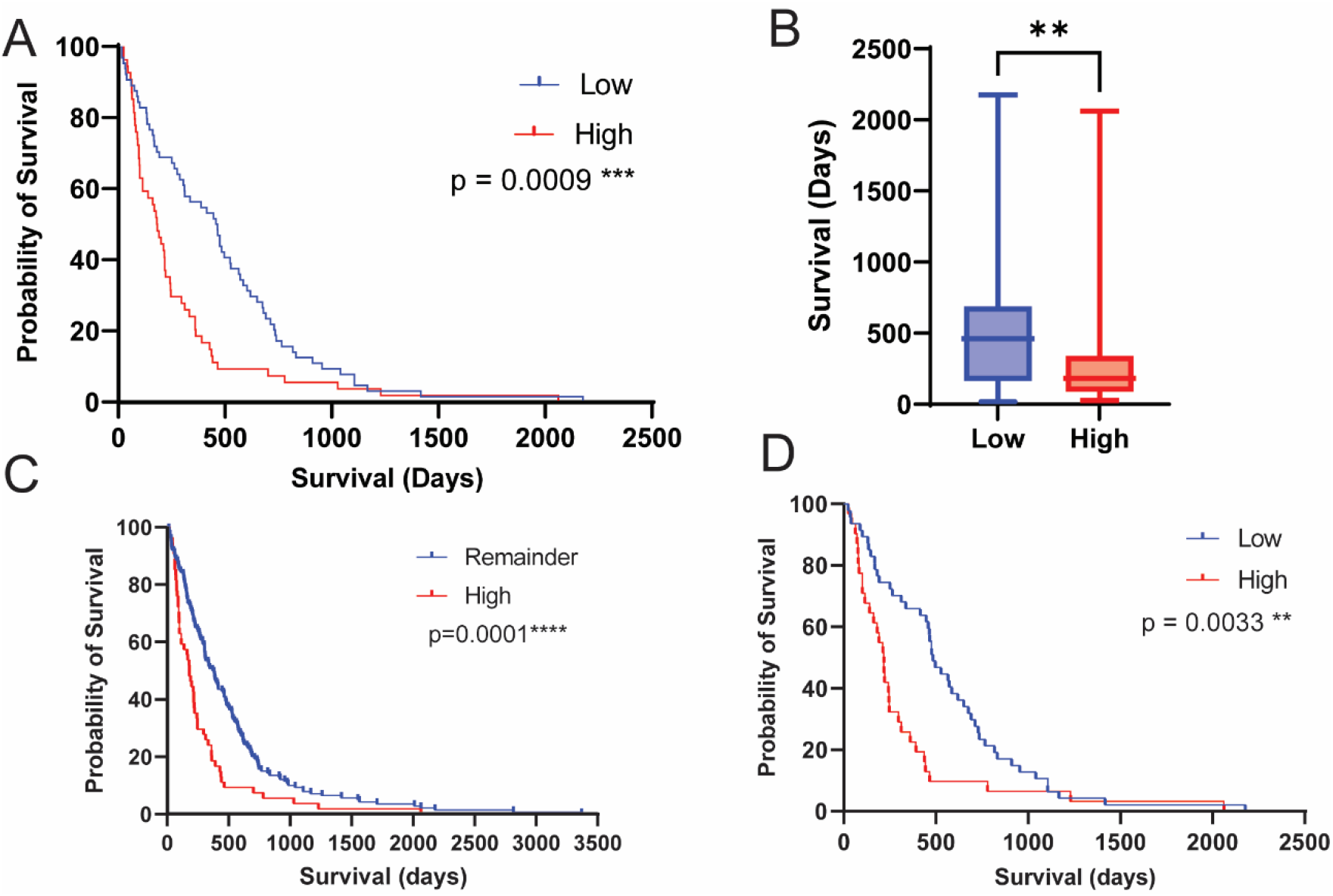
GCN2 Score is a predictor of prognosis. (**A, B**) Kaplan-Meier curve of GCN2 score in all mesothelioma cases, and box-whisker plot of survival in high (Upper two quintiles) and low (lower two quintiles) GCN2 scores. Median survivals: GCN2 High 181.5 Days, GCN2 Low 461.5 Days (**C**) High GCN2 group *vs* all other cases. Median survivals: GCN2 High 181.5 Days, Remainder 382.0 Days (**D**) High GCN2 Score *vs* Low GCN2 in epithelioid histology only. Median survival High 217.0 Days, Low 484.0 days.

### Multivariate Analysis

Using Cox regression in multivariate analysis, GCN2 was a significant indicator of survival, independent histology (See table 3). However, GCN2 scoring was not a significant predictor of survival independent of metastasis score (p=0.4643). The C-statistic value for GCN2 score was 0.690 (see Table 3), with histopathology (epi vs non-epi) performing marginally worse with a value of 0.657 Addition of the GCN2 score to histopathology logistic regression did not improve the C-statistic significantly.

**Table 3:** C-statistic values for 1 year survival.

| Model | C-statistic | $\Delta C$ (p-value) | CI 95% low | CI 95% high |
| --- | --- | --- | --- | --- |
| High GCN2 score | 0.690 |  | 0.6150 | 0.7657 |
| Histopathology | 0.657 |  | 0.5401 | 0.7022 |
| High GCN2 score<br>+ histopathology | 0.703 | 0.013 (ns) | 0.6096 | 0.7965 |

## Discussion

In this study we have examined the expression levels of GCN2 in malignant pleural mesothelioma (MPM) samples and correlated these levels with patient survival. Previous work had shown the CHOP levels predicted patient survival in MPM (25), however the work of Dalton *et al* had not determined the role of upstream signalling which may have contributed to those elevated CHOP levels. GCN2, being part of the Integrated Stress Response (ISR), may influence CHOP levels as much as PERK/Endoplasmic Reticulum stress. Furthermore, recent progression in pharmaceutical trials targeting ASS-1 dependency in MPM cases suggest a role for GCN2 in MPM.

One weakness of the study presented here is that we are unaware of the activation status of the GCN2 which have identified. Determining whether the higher expression of GCN2 correlates to increases in eIF2α levels, or a higher proportion of GCN2-autophosphorylation levels, as previously observed (30) would strengthen the association between GCN2 levels and CHOP. Additionally, it remains to be seen whether GCN2 overexpression is also responsible for ASS-1 level expression increases within this dataset.

In invasive bladder cancers, it has been demonstrated that ASS-1 loss promotes cancer cell survival through GCN2 expression (12), and that administration of ADI-PEG-20 causes apoptosis through GCN2/ATF4 mediated CHOP activation. Interestingly, ASS-1 protein and mRNA levels have recently been shown to be dependent on BRCA-associated protein 1 (*BAP1*) levels in MPM cells and tumours (31). Although exact percentages vary between 23%(32) and 49%(33), it has been shown in several studies that a significant proportion of MPMs have lower BAP1 levels, and germline mutations in the *BAP1* gene which lower protein levels and reduce nuclear localisation are associated with a predisposition to MPM (34). Through engineering MeT5A-*BAP1*^*w-/KO*^ cell lines which phenocopy BAP1-deficient cancers, Barnett *et al*. also showed that the BAP1 mediated increase in ASS-1 levels conferred greater resistance to ADI-PEG-20 (31) – which may in turn mean that these tumours do not have elevated GCN2 levels. In summary, we have shown that GCN2 levels are a useful prognostic biomarker for MPM survival. This may suggest that targeting GCN2 through small molecules, such as GCN2iB and others (23,35), may be a fruitful avenue for future therapeutic development.

## Data Availability

All data produced in the present study are available upon reasonable request to the authors

## Acknowledgements

We thank the Dundee Imaging Facility and the Data Analysis Group at the University of Dundee and those at the MesobanK for their help in this work. We thank the Tayside Biorepository for useful conversations and help with ethical and legal requirements. We thank Professor Stefan Marciniak for helpful discussion and feedback on the study. Material used in the Research Programme was obtained from Mesobank, a Research Tissue Bank supported by Asthma and Lung UK and The Victor Dahdaleh Foundation.

## Author’s Contributions

LTG conducted image scoring, data analysis and manuscript preparation. SB and NMK conducted slide staining and provided advice and training to LTG on scoring. NH and MN provided additional scoring data. GRM devised the experiments, conducted data analysis and also aided in manuscript and figure preparation.

### Ethics approval and consent to participate

All samples were collected and used under ethic and consented approval to participate in the study. All tumour samples obtained from MesobanK UK, Cambridge. Ethical approval for MesobanK was granted by the East of England-Cambridge Central Research Ethics Committee (18/EE/0161) on 9th August 2018.. This work uses data that has been provided by patients and collected by the NHS as part of their care and support. The data are collated, maintained and quality assured by the National Cancer Registration and Analysis Service, which is part of NHS Digital. Access to the data was facilitated by the UK Health Security Agency Office for Data Release

### Data availability

Blinded IHC data are available on request to the corresponding author.

### Competing Interests

We declare no competing interests.

### Funding information

LTG is funded through the Ninewells Cancer Campaign. GRM is funded by a Baxter Fellowship from the University of Dundee.

## Notes

### Competing Interest Statement

The authors have declared no competing interest.

### Author Declarations

Ethical approval for MesobanK was granted by the East of England - Cambridge Central Research Ethics Committee 18/EE/0161 on 9th August 2018

